# Expectation of Tactile Signals in Human Motor Cortex

**DOI:** 10.1101/2025.09.30.25336988

**Authors:** Natalya D. Shelchkova, Ali H. Alamri, Alexandriya M. X. Emonds, John E. Downey, Charles M. Greenspon

## Abstract

Somatosensory feedback and attention are critical for the execution of dexterous behaviors. Indeed, tactile and proprioceptive signals are sent to motor cortex such that they can be integrated into the motor plan while the expectation of feedback can shape motor responses. One downside to many of the studies that have found these interactions is that motor cortex has always been studied within the framework of a motor task, potentially squashing many of the smaller signals present and making others hard to disentangle. In this study we specifically investigate the presence of sensory expectations in human motor cortex in the absence of a motor task using two participants implanted with electrode arrays in both sensory and motor cortices, allowing us to identify signals corresponding to sensory expectation with fewer confounds. We found that the deployment of attention to individual fingers results in digit-specific activation of motor cortex while sensory cortex remains unperturbed. Moreover, we observed that, compared to an imagined movement task, the expectation signal in motor cortex existed in a distinct subspace that does not represent pure motor intent. Finally, we found that the expectation signals existed along a somatotopic axis that matched the somatotopic axis revealed by movement intent. This work highlights the fact that many signals are multiplexed in motor cortex.

## Introduction

Sensorimotor integration is fundamental to the execution of successful manual behaviors as motor cortex (MC) must incorporate sensory feedback to accurately update commands as new sensory information arises^1^. Indeed, it has been shown that motor cortex responds to tactile^2–6^ and proprioceptive^3–7^ manipulation. Furthermore, the removal of sensory and proprioceptive inputs impairs motor behavior^8^. Suppression of tactile inputs (e.g. at the cuneate nucleus or thalamus) results in decreased ability to coordinate dexterous behavior and use relevant tactile signals to perform motor behaviors^9^ while lesions to macaque somatosensory cortex (SC) impair dexterous hand control^10–12^. In addition, lesions in SC results in the inability to learn new motor behavior^13^, highlighting the importance of sensory inputs not only to the coordination of behavior but also in the acquisition of new motor skills.

Early work in macaques showed that the neural activity in MC differs when monkeys were cued to push or pull a lever in response to a perturbation prior to the execution of the movement^14^ and more recent work has shown that not only the direction but also the probability of perturbation can be decoded from activity in MC^15^. Although the presence of sensory signals in motor cortex is well established, it has always been investigated in conjunction with the preparation and execution of a motor task. As a result, it is difficult to disentangle the expectation of a stimulus occurring from the integration of this expectation into a subsequent motor plan. Understanding this distinction is crucial not only for our understanding of sensorimotor planning, but also for developing better brain-computer interfaces as these signals may exist in similar subspaces. Recent prosthetic research has demonstrated the benefits of incorporating artificial sensory feedback for robotic hand and arm control^16,17^, but has also shown that the delivery of this feedback can evoke activity in MC, inhibiting attempts to decode motor intent^18^. Consequently, there may be a similar overlap between sensory expectation and motor intent or planning and this could influence attempts to decode motor intent.

In this study we decouple sensory expectation from movement planning by having two human participants perform a modified spatial attention task in which they were cued towards ICMS delivered to different fingers on each trial to induce sensory expectation. We find that while expectation did not modulate SC, substantial changes were observed in MC. Moreover, we find that the expectation signals in MC reflect which finger the participant was attending to and that these signals are distributed in a somatotopic manner. Finally, we demonstrate that, while similar, the expectation signals and motor intent signals exist in distinct subspaces. These findings reinforce previous work showing that somatotopy plays a distinct role in MC at the individual finger level by observing similar somatotopic tuning for both sensory expectation and motor intent signals and demonstrates the added layers of context that are present in MC.

## Results

To assess whether sensory expectations are present in MC we asked two participants implanted with Blackrock NeuroPort arrays in the motor and somatosensory cortices (**Figure 1A**) to perform a modified Posner task^19^. We delivered intracortical microstimulation (ICMS) through two individual electrodes of the sensory arrays (**Figure 1B**) to elicit sensations on individual digits (**Figure 1C**), the location of which the participants were cued toward on each trial (**Figure 1D**). *Before testing*, we computed the amplitude range between which the participant could detect 5% and 95% of trials so that a psychometric function could be created for each electrode. *During testing*, a plurality of trials (30%) were at the estimated detection threshold amplitude (50^th^ percentile), 20% of trials were at the 37.5^th^ and 62.5^th^ percentiles, and 10% were at the 12.5^th^ and 87.5^th^ percentiles. As a control, 10% of trials for each electrode were catch trials where no stimulation was delivered. On each trial the participant was cued to which digit a stimulus would occur on and was asked to report if they detected a stimulus on any digit. On 90% of the trials the cued and stimulated digits were congruent, which we refer to as valid trials, and in 10% the cued and stimulated digits were incongruent, which we refer to as invalid trials.

**Figure 1.**
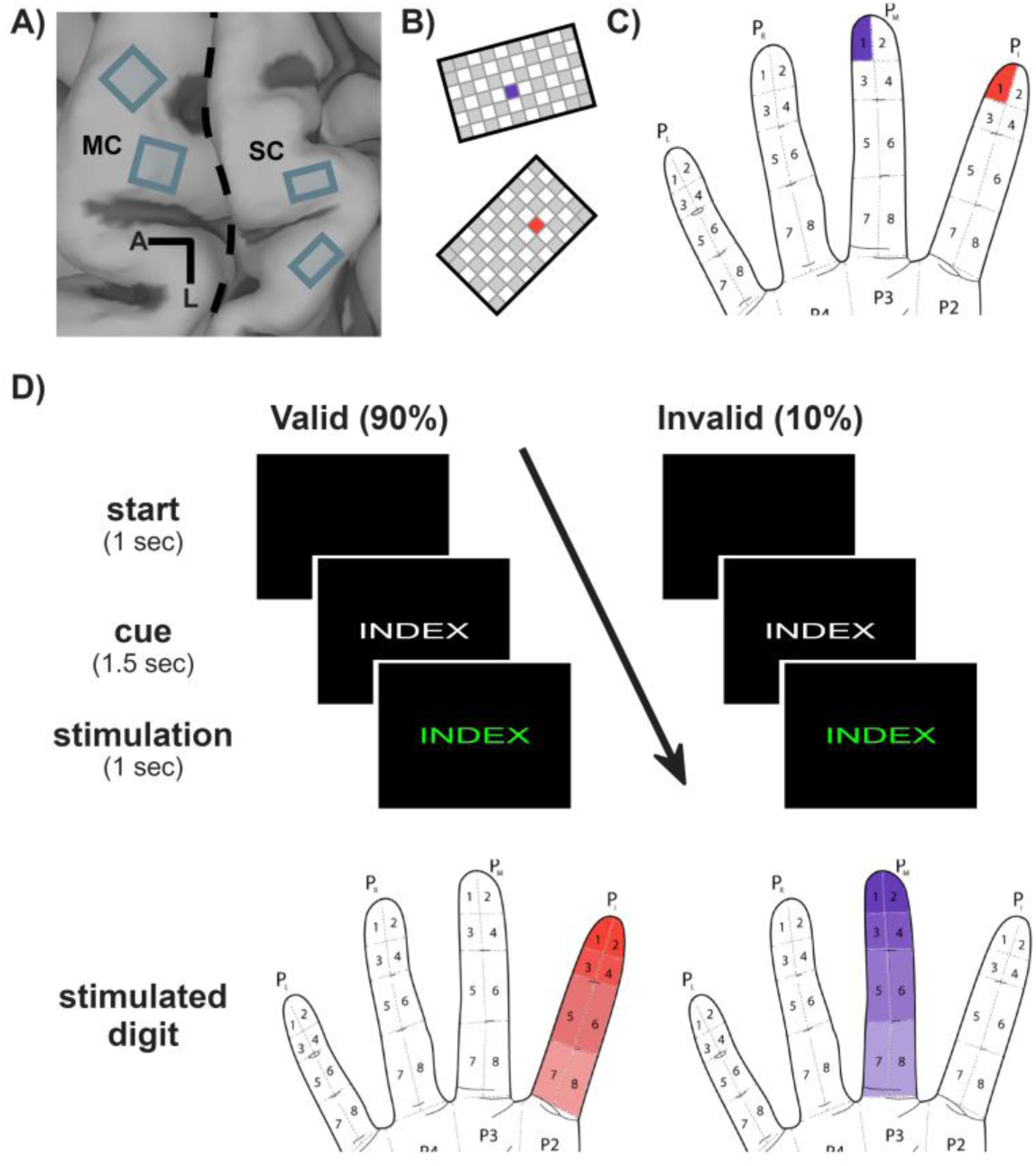
Tactile Posner task. **A)** MRI of participant C1’s brain with overlayed NeuroPort arrays implanted into motor (MC) and somatosensory cortex (SC). **B)** Electrode arrangement on the sensory arrays, white squares indicate wired electrodes while gray squares are unwired electrodes. The purple electrode elicits a sensation on the middle finger when stimulated, while the red electrode results in a sensation on the index finger. **C)** The location of the ICMS-evoked percept, as reported by the participant, when stimulating through the purple and red electrodes in B. **D)** Posner task layout. On each trial the participant was cued to which digit they must attend to. After the cue, stimulation was delivered to one of two electrodes. The text color changed to green at the start of stimulation and back to white when stimulation ended. In the valid condition, the same digit that is cued is stimulated, in the invalid condition, another digit is stimulated.

A total of 14 sessions were performed (n = 12 and 2 for participants C1 and C2, respectively). To confirm that our participants were sufficiently attending to the task, we computed the false alarm rate for each electrode and found that only 10/28 sessions had a false alarm rate above 25% (**Supplementary Figure 1A**). These sessions were not removed from analysis as the results did not substantially differ from sessions with lower false alarm rates. To first compare task performance with more classic implementations of the Posner task performed with monkeys^20^, we computed psychometric functions for each electrode and compared detection rates for valid and invalid trials (**Supplementary Figure 1B**). While some sessions showed signs of attentional modulation, where detection rates were lower when the participant was miscued (3/24 sessions, permutation test: p < 0.05), others showed no effect (12/24) or even the opposite response where invalid trials resulted in better detection compared to valid trials (13/24). This implies that while the participant was performing the task, either their ability to deploy their attention was less granular than that of monkeys performing visual tasks, that the shift from a discrimination task to a detection task reduced the task’s sensitivity to detecting attentional shifts, or that the orienting response overpowers the expected attentional effect. The majority of the results presented below reflect participant C1, while several experiments were replicated in participant C2 who’s results are shown in **Supplementary Figure 2**.

### Sensory expectation is poorly represented at the single electrode level

We first investigated whether the presentation of a cue resulting in deployment of spatial attention and thus sensory expectation modulates motor or somatosensory cortex at the single electrode level. Using a 1-way ANOVA to assess modulation of the multi-unit activity (MUA), we found that both motor and sensory MUA were modulated during each period of the task. Motor MUA was modulated approximately 500 ms after cue onset in both excitatory (**Figure 2A left, Supplementary Figure 2A left/middle**) and inhibitory (**Figure 2A middle**) manners (32.6% ± 16.9% excitatory and 7.1% ± 4.1% inhibitory responses across sessions, Dunnet’s post hoc, p < 0.01). After stimulation, differences in response were observed, reflecting electrode specific activation of MC^18^. The effects on the sensory arrays were more subtle, with very few channels demonstrating modulation to the cue (3.5% ± 2.7% excitatory, 0.8% ± 1.6% inhibitory responses across session, Dunnet’s post hoc, p < 0.01) likely in part due to low baseline firing rates, though significant modulation was observed immediately after the ICMS train (**Figure 2A right, Supplementary Figure 2A right**). Note that recordings during stimulation on the sensory arrays are unreliable due to stimulus artifacts and so are not considered here.

**Figure 2.**
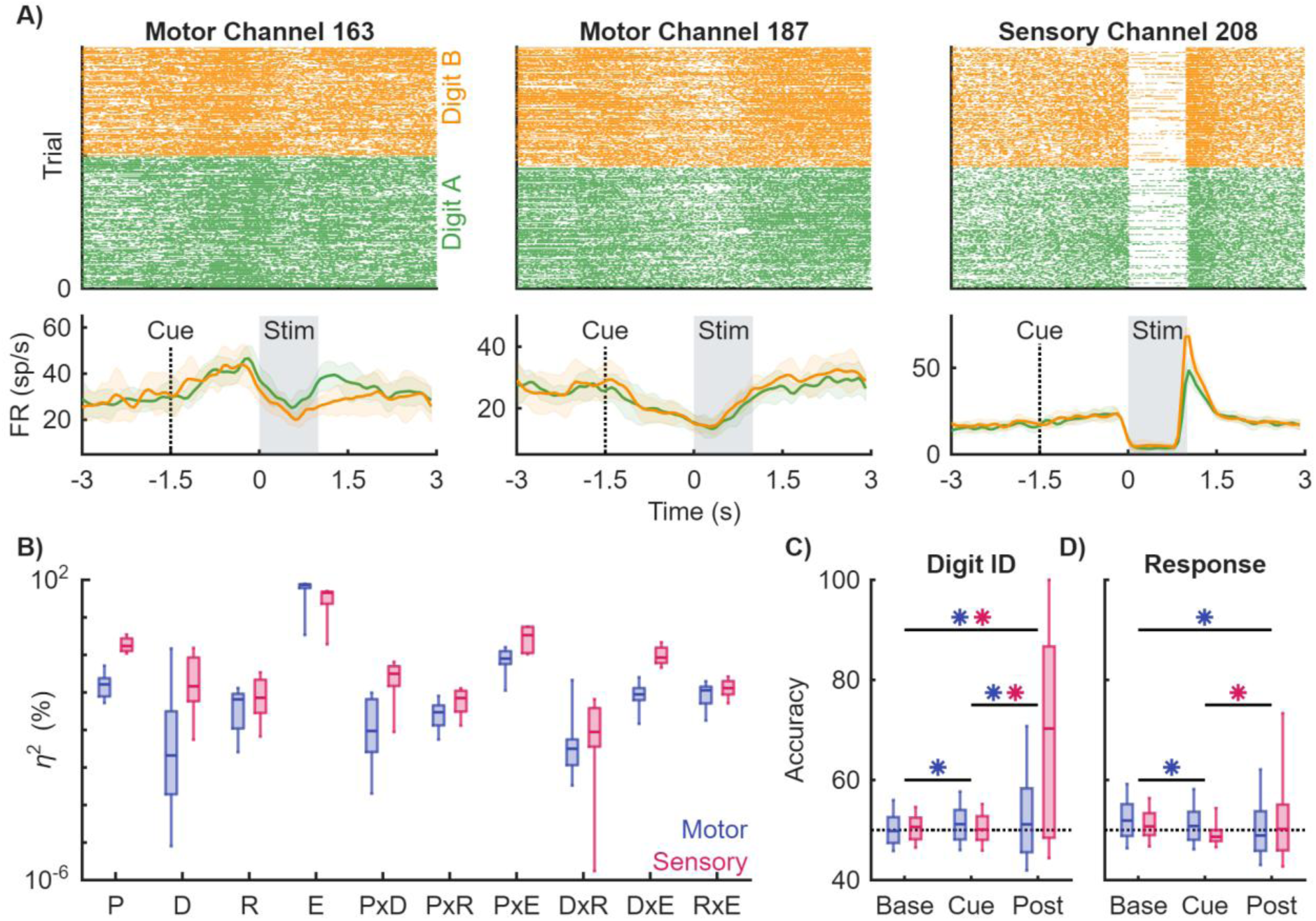
Neural correlates of sensory expectation in motor and somatosensory cortex. **A)** Rasters (top) and post-stimulus time histograms (bottom) of neural responses throughout the trial. Green and orange indicate cues to different digits. **B)** Variance explained by the period (P) (baseline, cue, post-stim), the digit cued (D), participant’s response (R), and electrode (E). Interaction terms are denoted with a x. **C)** Decoding accuracy for digit ID split by motor (blue) and sensory (pink) arrays. Dashed line indicated chance classification performance. Boxes indicate 5^th^, 25^th^, 50^th^, 75^th^, and 95^th^ percentiles of variance explained across sessions and channels. Asterisks denote a significant difference in performance determined by a 1-way ANOVA (p<0.001, Tukey-Kraner post hoc). **D)** Same as C but for participant response. Data from participant C1.

To quantify the degree of modulation by task period and cued digit, we performed a 4-way ANOVA on the firing rate across each period (baseline, cue, and post-stimulation) conditioned by the cued digit, the participants response, and the electrode (**Figure 2B, Supplementary Figure 2B**). We found that, across sessions, electrodes accounted for most of the variability observed (eta^2^, η^2^: 60.73% ± 27.36% motor, 35.71% ± 22.66% sensory). For the task conditions: period had the largest impact (η^2^: 0.19% ± 0.14% motor, 2.01% ± 1.06% sensory) followed by the cued digit (0.15% ± 0.47% motor, 0.46% ± 0.72% sensory) and finally the participant’s response (0.06% ± 0.05% motor, 0.12% ± 0.15% sensory). Additionally, sensory arrays showed the largest interaction between period and digit during the post-stimulation period because much of the post-stimulation activity in the sensory arrays was driven by the injection of electrical current from the ICMS^21^.

We next sought to quantify the ability of single electrodes to classify task conditions: cued digit and the participants response. Using the firing rates during each period, we used linear discriminant analysis (LDA) to classify each condition at each period for the motor and sensory arrays separately (**Figure 2C, Supplementary Figure 2C**). All trials were included for classification during the baseline and cue periods, as the participant was unaware of the valid/invalid condition, while only the valid trials at the detection threshold amplitude were included for the post-stimulation period. Classification of the cued digit was at chance for both motor and sensory arrays during baseline and cue (49.8% ± 5.7% motor baseline, 50.1% ± 5.2% sensory baseline, 51.0% ± 6.0% motor cue, 49.9% ± 5.3% sensory cue), implying that information about sensory expectation is encoded at the single electrode level before stimulation. However, classification performance significantly increased after stimulation for both motor and sensory electrodes (52.6% ± 10.6% motor, 69.5% ± 20.3% sensory). This increase in decoding accuracy is likely driven by the activation of both regions following stimulation^18,21^. We repeated the same analysis instead attempting to classify the participant’s response and found that we were unable to do so throughout all periods of the trial and for both motor and sensory arrays (**Figure 2D, Supplementary Figure 2D**).

### Sensory expectation is distributed across the population

To determine if task condition signals were more robustly embedded in the population response, we attempted to classify the cued digit or the participant’s response using the firing rates from an increasing number of electrodes (**Supplementary Figure 3A,B**). For both motor and sensory arrays, slight improvements in classification performance of digit identity during the cue period were observed as the number of included electrodes increased, however these improvements were modest (single channel -> multi-channel: 52% - > 57% and 50% -> 53% for the motor and sensory arrays, respectively). As with the single electrode analysis, classification during the post-stimulus period dramatically increased – reaching 66% in motor and ∼100% in sensory – again likely due to ICMS-evoked activity. When attempting to classify the participant’s response, we found increases in performance from chance when including more electrodes. Surprisingly, we were also able predict the participant’s response based on the baseline activity above chance (55% and 57% motor and sensory respectively), implying a bias signal in both MC and SC (**Supplementary Figure 3C,D**).

Given this putative success, we investigated if dimensionality reduction would allow for better isolation of underlying neural patterns that might represent task information. To assess this, we performed PCA on the condition-averaged data for cued digit identity and participant response separately and then projected the raw firing rates onto the top 10 PCs. Examining the neural trajectories in PC space conditioned on digit identity for the motor arrays, we found that after the cue, the trajectories for the individual digits began to separate and that this separation was maintained after stimulation (**Figure 3A, Supplementary Figure 2E**). Given the apparent low dimensional nature of this signal, we performed LDA classification after projecting onto a subset of ascending PCs. We found that asymptotic performance could be reached within 2-3 PCs for the motor array (50.5% ± 3.3% baseline, 59.2% ± 6.6% cue, 65.4% ± 8.0% post-stimulation using up to PC3) (**Figure 3B, Supplementary Figure 2F**) and that the vast majority of performance gain was from an individual PC, confirming the low-dimensionality of the signal. Looking at the neural trajectories of the sensory arrays, we found no separation for the individual digits and classification performance was largely driven by stimulation (**Figure 3C**) such that digit identity could only be decoded after stimulation (**Figure 3D**). Moreover, due to the large variance induced by ICMS, asymptotic classification performance was achieved with the first PC alone (50.9% ± 0.9% baseline, 51.3% ± 3.3% cue, 69.5% ± 8.0% post-stimulation). These results were consistent across test sessions (**Figure 3E, Supplementary Figure 2G**), regardless of which pair of fingers was tested.

**Figure 3.**
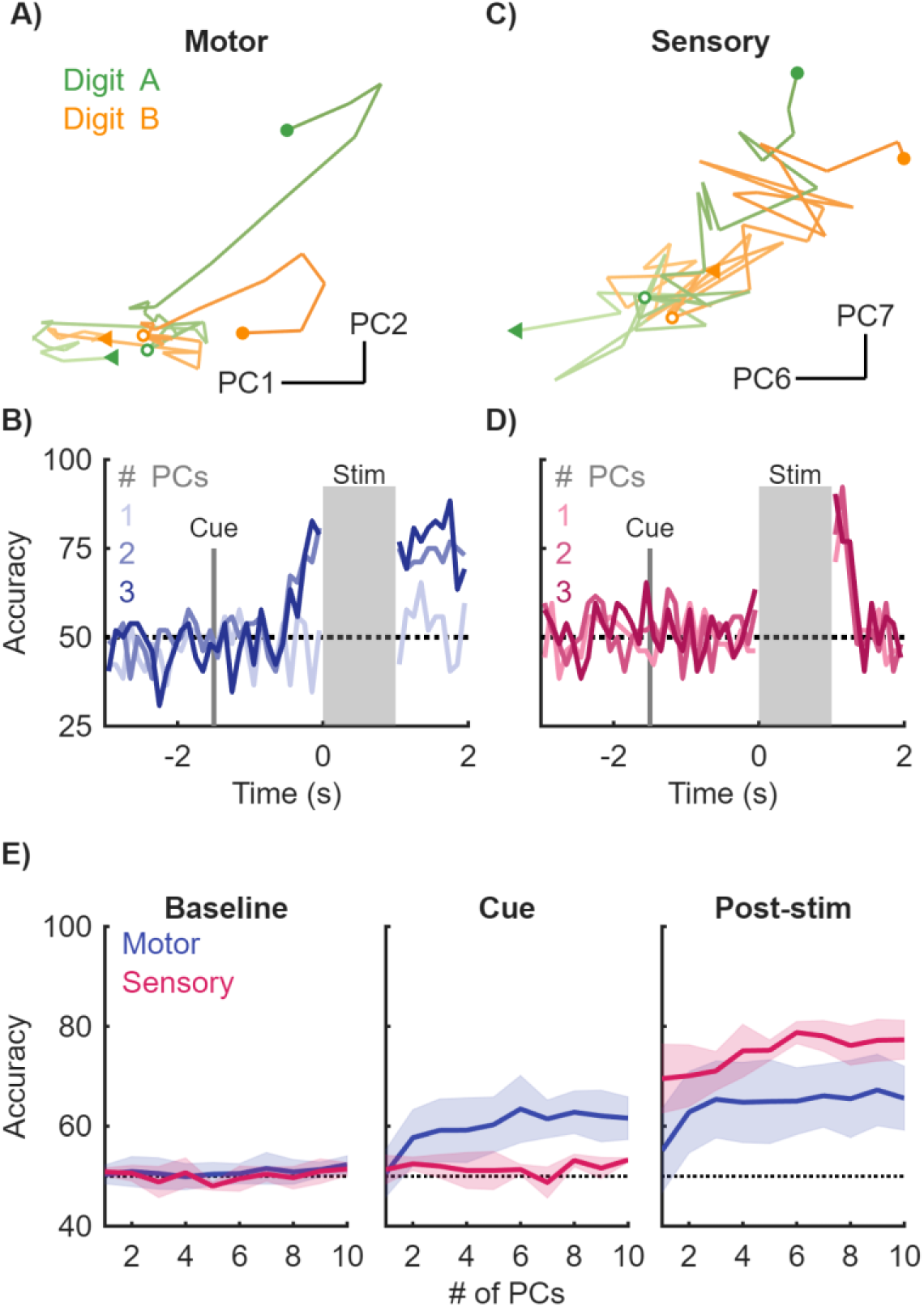
Digit identity can be decoded from population responses. **A)** Activity from the motor array projected into PC space. Green and orange traces represent the two digits cued. The start of the trial is denoted by a triangle, the cue onset is denoted by an open circle, and the start of stimulation is denoted by a filled in circle. **B)** Decoding of digit identity from the motor array using the first 3 PCs. Dashed line indicates chance performance. **C)** The same as A but for the sensory arrays. **D)** The same as B but for the sensory array. **E)** Average decoding performance across sessions as a function of the number of PCs used for the motor (blue) and sensory (pink) arrays. Performance was grouped into baseline, cue, and post-stimulation epochs by averaging performance during a 1 second interval for baseline and 500ms period for cue and post stimulation. Mean and STD indicated by line and shaded area. Dashed line indicates chance. Data from participant C1.

Using the same analysis to decode the participant’s response (**Supplementary Figure 4**), we saw little separation between trials in which the participant reported detecting the stimulus or not, with the greatest separation instead occurring at the beginning of the trial (**Supplementary Figure 4A, Supplementary Figure 2H**). The dimensionality of this signal was also quite low, with asymptotic performance achieved using 2-3 PCs (**Supplementary Figure 4B, Supplementary Figure 2I**). Additionally, our ability to decode the participant’s response was slightly above chance during the baseline period, and we had marginal increases in performance for the cue and post-stimulation periods (53.0% ± 3.1%, 56.2% ± 3.9%, 56.5% ± 5.6% for baseline, cue, and post-stimulation respectively using up to PC3). When analyzing the activity on the sensory arrays, the neural trajectories began separating with higher PCs (**Supplementary Figure 4C**), though decoding performance remained at slightly above chance level throughout the trial (**Supplementary Figure 4D**) even when all 10PCs were used (54.1% ± 2.8%, 53.7% ± 4.0%, 56.6% ± 5.2% for baseline, cue, and post-stimulation respectively, **Supplementary Figure 4E, Supplementary Figure 2J**).

### Sensory expectation is distinct from movement intention

As stated in the introduction, a common confound in these experiments is the co-occurrence of sensory expectations with motor planning. Although our task explicitly did not contain a motor component and we instructed our participant not to imagine any movements, it is of course possible that the participant incorporated some movement plan that would influence our results. To address this, in a single experimental session we performed both a 4-finger variant of the previous modified Posner task as well as a finger movement task such that the signals could be compared. The motor task followed the same general layout of the Posner task with start, cue, and movement phases to ensure parity between the tasks. We first examined if the sensory expectation results when performed with 4 fingers matched those with 2 and found that both the individual electrode (**Supplementary Figure 5**) and population analyses (**Supplementary Figure 6**) were consistent between variants.

Computing PCA on the combined datasets and comparing the trajectories between the tasks (**Figure 4A,B**), we found that there were some similarities in that the thumb was consistently distinct from the other digits for both tasks. Crucially, while the trajectories were qualitatively similar, projecting each dataset into the others PC space revealed drastically different locations in the space (**Supplementary Figure 7**). Next, we attempted to use LDA across datasets to determine if, despite the differences, some residual information remained. Thus, we first fit an LDA classifier on each dataset and then attempted to classify the other dataset using the same classifier (**Figure 4C, Supplementary Figure 8**). We found that the classifier trained on the movement data was excellent at classifying which finger the participant was attempting to move (∼95%, p<0.001, shuffled control) but only relatively poorly captured which finger the participant was attending to in the Posner task (34.5%, p<0.001, shuffled control). Conversely, the classifier trained on the sensory expectation dataset was able to predict movement and Posner tasks with similar performance (61.9% and 51.0%, respectively, p<0.001, shuffled control). This confirms both that the expectation signals are not movement intent and that the movement task contains both elements of expectation and movement planning, though these exist in two distinct subspaces. Finally, having observed this apparent similarity and given our previous findings on the somatotopy of ICMS-evoked activity in MC^18^, we sought to determine if this might be reflected in the expectation signal. Consequently, we computed the average digit preference for each electrode (from thumb to ring) for both motor and expectation tasks (**Figure 4D**). We observed a trend across the medio-lateral axis for thumb-preferring to ring-preferring electrodes for both tasks and the electrode-specific preferences were highly correlated between tasks (**Figure 4E**) suggesting that the somatotopic tuning was shared across movement and expectation tasks.

**Figure 4.**
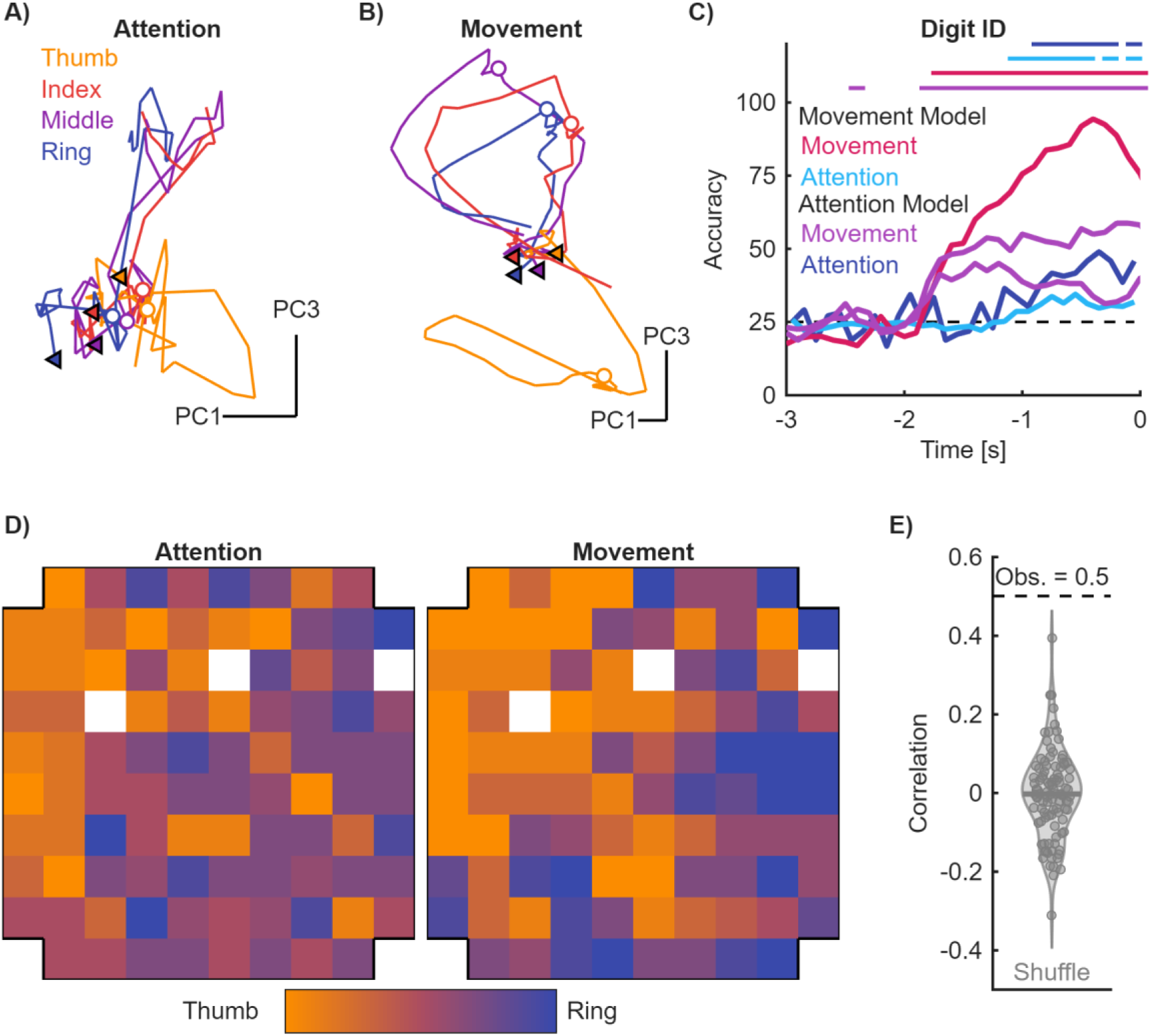
Movement and sensory expectation. **A)** Activity from the motor array during the expectation task projected into PC space. Triangles indicate the start of trial and open circles indicate cue onset. Different colors denote different digits. **B)** Activity from the motor array during the movement task projected into PC space. Triangles indicate the start of the cue period and open circles indicate start of flexion. **C)** Average decoding performance for digit identity using expectation and movement models. Neural data during the expectation task was used to decode digit identity using a model trained on the expectation data (blue) or the movement data during the preparation period (light blue). Neural data during the movement task was used to decode digit identity using a model trained on the movement data (pink) and on the expectation data (purple). Horizontal lines indicate significant performance compared to shuffled models (p ≤ 0.05). **D)** Heatmaps of the digit preference of each channel in the motor array during the expectation and movement tasks. **E)** The observed correlation (dashed line) between the heatmaps in D compared to shuffled correlations (gray). Data from participant C1.

## Discussion

### Attention weakly modulates SC

In this study we investigated the role that sensory expectation and attention have on motor and sensory cortical activity. When examining the activity in somatosensory cortex, we found little to no modulation to the digit cued across any of the channels. This is unsurprising however as signatures of attention in somatosensory cortex are more prevalent in secondary somatosensory cortex (S2) of both humans^22,23^ and monkeys^24–31^, though some have reported activation in primary somatosensory cortex as well^32^. Importantly, in the few studies that observed modulation in S1, it was greatest in layer I/II and weakest in deeper layers^28^. This is relevant because our arrays target layer IV and so may be why we did not observe any modulation here.

Moreover, the task design, while inspired by the Posner task and variants performed in macaques, was sufficiently different that inconsistent results are not unreasonable. First, the fact that we performed a detection task instead of a discrimination task meant that the baseline firing rate was very low, potentially creating a floor effect. This decision was made both because extended ICMS can cause adaptation^33^ and because closely space ICMS trains causing substantial excitation^34^, either of which would have made reliably delivering stimulation in the peri-threshold range infeasible. Second, macaques are often *extensively* trained to the point of overtraining whereas the human participant was exposed to this task for the first time with no training beyond existing detection threshold tasks and sessions were interspersed among many other tasks. Third, it’s unclear to what extent the participant was truly focusing on one finger or the other or if the ability to spatially attend is distinct between tactile and vision. Finally, it must be acknowledged that only one participant was used in this study due to the difficulty of scheduling BCI experiments. Nevertheless, it must also be noted that even if the results were different with a second participant, the varying nature of the injuries would make interpretation difficult.

### Sensory expectation signatures were most prominent in MC

Despite the sensory nature of the task, motor cortex was significantly more modulated by expectation than sensory cortex. While this might be surprising given that the participant was performing a tactile attention task with no motor response, the modulation of MC is not unprecedented in attention tasks^22,30^, though these tasks included motor components. Given that motor cortex receives input from somatosensory cortex^35–43^, is known to respond to tactile inputs^3–6^ and even encodes predictions of sensory perturbations^44^, we interpret this attentional modulation as a signature of sensory expectation. Previous work in vision has postulated a ‘premotor theory of attention’^45,46^ which suggests that the same brain areas involved in allocating attention are also involved in movement preparation. Within the frontal eye fields (FEF) neurons have been identified that respond to allocation of attention, preparation of saccade movements, and combinations of both^47,48^. These have been determined as distinct classes of neurons, whose activity can modulate each other but also other higher order regions including parietal areas^49^.

More recent efforts in humans using ECoG arrays, specifically for speech prosthetics, have highlighted the notion of shared representations in motor areas^50^. Specifically, electrodes whose temporal activation profiles differed between reading, listening, and attempted speech were observed. When investigating if the signal we were decoding from the motor arrays during the expectation task was in fact movement intention, we found that, while some information about movement intent could be decoded, the signal was significantly weaker during the expectation task. If attending to a digit in expectation of a sensation and moving it were indeed the same, we would expect equivalent performance in both conditions, especially as our participant was not making overt movements, but instead imagining them due to the nature of their injury. These results instead imply that attention is deployed for both tasks, but then ultimately used for different purposes.

### Somatotopy and context are prevalent in MC

Recent studies of the precentral gyrus have shown a coarse somatotopic representation^51–53^. However, we previously showed a comparatively fine somatotopic representation for the movement of individual fingers^18^ where a clear gradient was shown on the scale of millimeters, a result we have replicated here. Notably, the same participant was used in both this paper and in those that showed coarse somatotopy. Moreover, we find, for the first time, that the somatotopy extends beyond movement intent and sensory feedback but also to more cognitive signals such as attention and expectation. These patterns have likely gone unnoticed in primate studies due to a lack of focus on the movement of individual fingers and in human MRI studies due to poor spatial resolution. The main implication of these results is that cognitive signals can be decoded from motor cortex and that they are likely to be multiplexed with motor intention. In conjunction with our previous findings that ICMS of SC can evoke activity in MC^18^, this adds another layer of complexity to the inputs that are likely to influence motor decoding. While many efforts have necessarily focused on relatively simple cursor and robotic arm movement tasks constrained to the laboratory, real world usage is likely to introduce more complex variables that decoders will need to account for and be invariant to in order to ensure robustness and flexibility.

## Methods

### Participants

This study was conducted under an Investigational Device Exemption from the U.S. Food and Drug Administration and approved by the Institutional Review Board at the University of Chicago. The clinical trial at the University of Chicago is registered at ClinicalTrials.gov (NCT01894802). Informed consent was obtained before any study procedures were conducted. Participant C1 (male, m), 55-60 years old at the time of implant, presented with a C4-level ASIA D spinal cord injury (SCI) that occurred 35 years prior to implant. Participant C2 (male, m), 60-65 years old at time of implant, presented with a C4-level ASIA D SCI, along with a right brachial plexopathy, that occurred 4 years prior to implant.

### Cortical implants and stimulation

Four NeuroPort microelectrode arrays (Blackrock Neurotech, Salt Lake City, UT, USA) were implanted in participant C1 and C2, two in the hand and arm representation in motor cortex and two in the hand representation of Brodmann’s area 1^54^. The motor arrays are arranged in a 10 x 10 grid with the corners disconnected for a total of 96 channels. The sensory arrays are 6 x 8 with 32 electrodes arranged in a checkerboard pattern. ICMS was delivered via a CereStim 96 (Blackrock Neurotech). Each stimulating pulse consisted of a 200-µs cathodic phase followed by a half-amplitude 400-µs anodic phase (to maintain charge balance), the two phases separated by 100 µs. A mixture of custom analogue headstages and Cereplex ES headstages (Blackrock Neurotech) were used to stimulate and record signals.

#### Detection Thresholds

To characterize the detection sensitivity of each stimulation channel, we employed a two-stage psychophysical procedure: an initial **unconstrained staircase** phase followed by a **constrained method of constant stimuli**.

##### Stage 1: Unconstrained Staircase Mapping

In the first stage, we estimated the dynamic range of detection for each channel using an adaptive linear sweep of stimulus amplitude. Stimulation began at the minimum amplitude and increased in fixed, 10 µA steps until the participant successfully detected five sequential trials, establishing the upper detection boundary. The amplitude was then decreased in the same stepwise manner to identify the lower bound of non-detection. Based on these bounds, a set of 5 linearly spaced amplitudes was selected to span the full detection range. Each amplitude was presented multiple times in pseudorandom order using a 2-alternative forced choice (AFC) paradigm where participants had to report in which of the two stages the stimulus was presented. These responses were used to fit an initial logistic psychometric function, from which we extracted the detection threshold (DT50)—the amplitude at which detection occurred on 50% of trials—and the inter-quartile range (IQR), defined as the amplitude range over which detection probability increased from 25% to 75%.

##### Stage 2: Constrained Method of Constant Stimuli

To refine the psychometric curve and improve the reliability of threshold estimates, we conducted a second stage focused on amplitudes near the DT50. A new set of fixed amplitudes was selected, centered on the DT50 and spanning the IQR. These amplitudes were presented in pseudorandom order, with each level repeated 2-12 times to increase sampling density in the transition region of the psychometric function. The refined detection data were fit with a cumulative logistic function, yielding more precise estimates of DT50 and IQR for each channel. This two-stage procedure allowed for efficient yet accurate characterization of perceptual sensitivity across stimulation sites.

#### Attention Task

The participant performed a modified Posner task which used intracortical microstimulation to target the digits. At the start of each trial a cue was presented on the screen in white text to indicate which digit the participant must attend to. After 1.5 seconds, the cue would turn green indicating the potential onset of stimulation. At the end of the stimulation period, the participant reported if they felt a sensation on any digit, regardless of the cue.

On 90% of the trials, stimulation occurred through a channel that elicited sensation on the cued digit, referred to as valid trials. The other 10%, stimulation would occur on the other channel, referred to as invalid trials. Additionally, 10% of the trials for each digit were catch trials where no stimulation occurred and were used to compute the false alarm rate for that digit. Stimulation amplitudes were determined using the psychometric curves computed from the detection threshold experiment. 30% of the trials used the DT50 stimulation amplitude for that channel, 40% of the trials were at a stimulation amplitude that was 0.75 * IQR above (20%) and below (20%) threshold, and 20% of the trials were at a stimulation amplitude that was 0.25 * IQR above (10%) and below (10%) threshold.

#### Movement Control Task

To control for the possible confound that sensory expectation in motor cortex is movement intent, the participant performed a digit movement task where they were instructed to observe and attempt to follow the movements of a virtual hand on the TV screen. On each trial, the participant was given an audio cue indicating which digit (thumb, index, middle, or ring) would be moved on that trial. After 1.5 seconds, a tone indicated the start of flexing the digit and a second tone, occurring 1 second after the first, indicated a return to rest position.

#### Linear Discriminant Analysis (LDA)

Single channel decoding of digit ID and participant response was performed using LDA. The average firing rate was computed for each channel during the baseline, cue, and post-stimulation period using a 1 second window. For each channel and period we used the firing rate for that period to predict either the cued digit or participant’s response using the *fitcdiscr* function in MATLAB. To ensure class balancing due to uneven participant responses, we used bootstrapping and repeated the classification 100 times, balancing the classes with each repetition. While this was broadly unnecessary for classifying the cued digit, it was done in the same manner for rigor. The decoding performance was computed as the average of these 100 bootstraps. To ensure that the decoder was not overfitted, each bootstrap used 5-fold cross-validation using the *cvpartition* function in MATLAB.

#### Principle Component Analysis (PCA)

For each channel, we computed the average firing rate binned into 100 ms bins from 1.5 seconds before the cue until 1 second after stimulation ended. Data from the stimulation interval was removed as we could not confidently separate the ICMS-evoked responses from those modulated by the task (0-1.05 seconds after stimulus start). PCA was performed on the condition averaged firing rates, split by motor and sensory arrays. Data from individual trials were then projected onto the first 1-10 PCs. LDA as described above was used on the projected PC scores.

For the cross-projected expectation and movement control data sets, PCA was performed on each dataset separately, using the methods described above. Raw data was mean subtracted and then projected onto the coefficients from either dataset. LDA was again used on the PC scores for classification.

## Data Availability

Code for data processing and analysis can be found on GitHub
(https://github.com/CorticalBionics/SensoryExpectationsMotorCortex). Deidentified data generated in this study have been
deposited in the Data Archive BRAIN Initiative repository (https://dabi.loni.usc.edu/dsi/UH3NS107714)

https://github.com/CorticalBionics/SensoryExpectationsMotorCortex

## Author Contributions

CMG and AHA conceived of the experiments. AHA and AMXE collected the data. NDS, AHA, and AMXE performed the analysis. NDS and CMG prepared the manuscript. CMG and JED secured funding. All authors edited the manuscript.

## Acknowledgments

We would like to thank the participants for their generous contribution to the advancement of science. The work at the University of Chicago was supported by the National Institute of Neurological Disorders and Stroke of the National Institutes of Health under Award Numbers UH3 NS107714, R35 NS122333, and R01 NS130302, all of which were developed with significant support from Sliman Bensmaia who we would like to thank. We would also like to acknowledge the members of the Cortical Bionics Research Group for their contributions and feedback.

## Supplementary Information

**Supplementary Figure 1.**
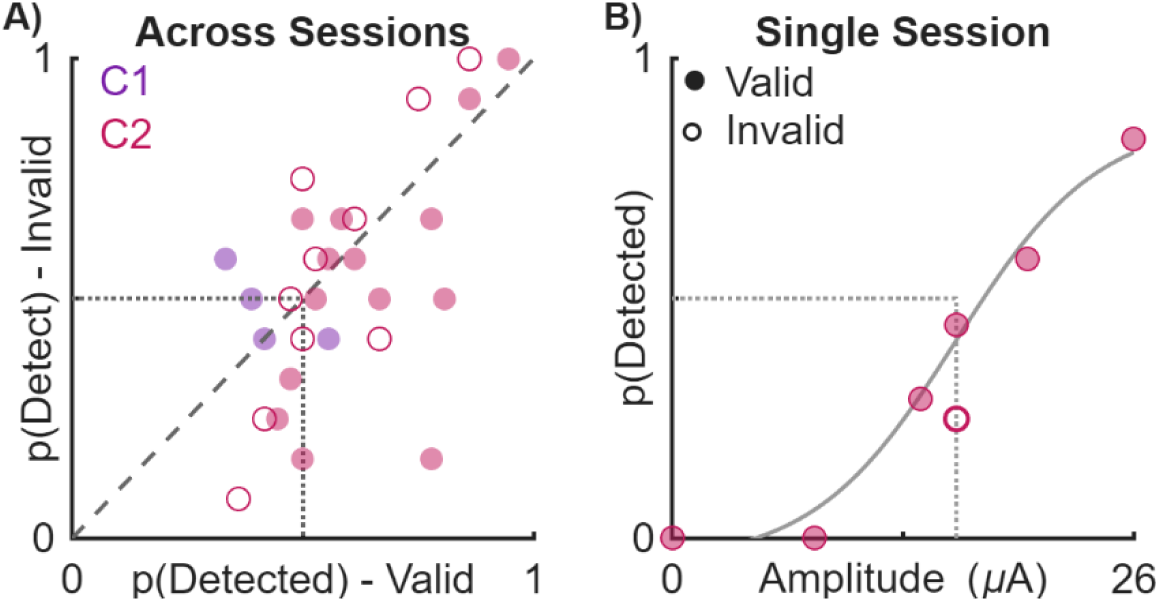
Performance during modified Posner task. **A)** Modulating attention during the modified Posner task had minimal impact of task performance. Each dot represents the performance for one digit and unfilled dots indicate those with high alarm rates (>= 25%). **B)** Example psychometric function for a channel showing the probability of detecting if a stimulus was present or not based on its amplitude. Filled circles indicate performance when a valid cue was presented while the open circle indicates performance for an invalid cue.

**Supplementary Figure 2.**
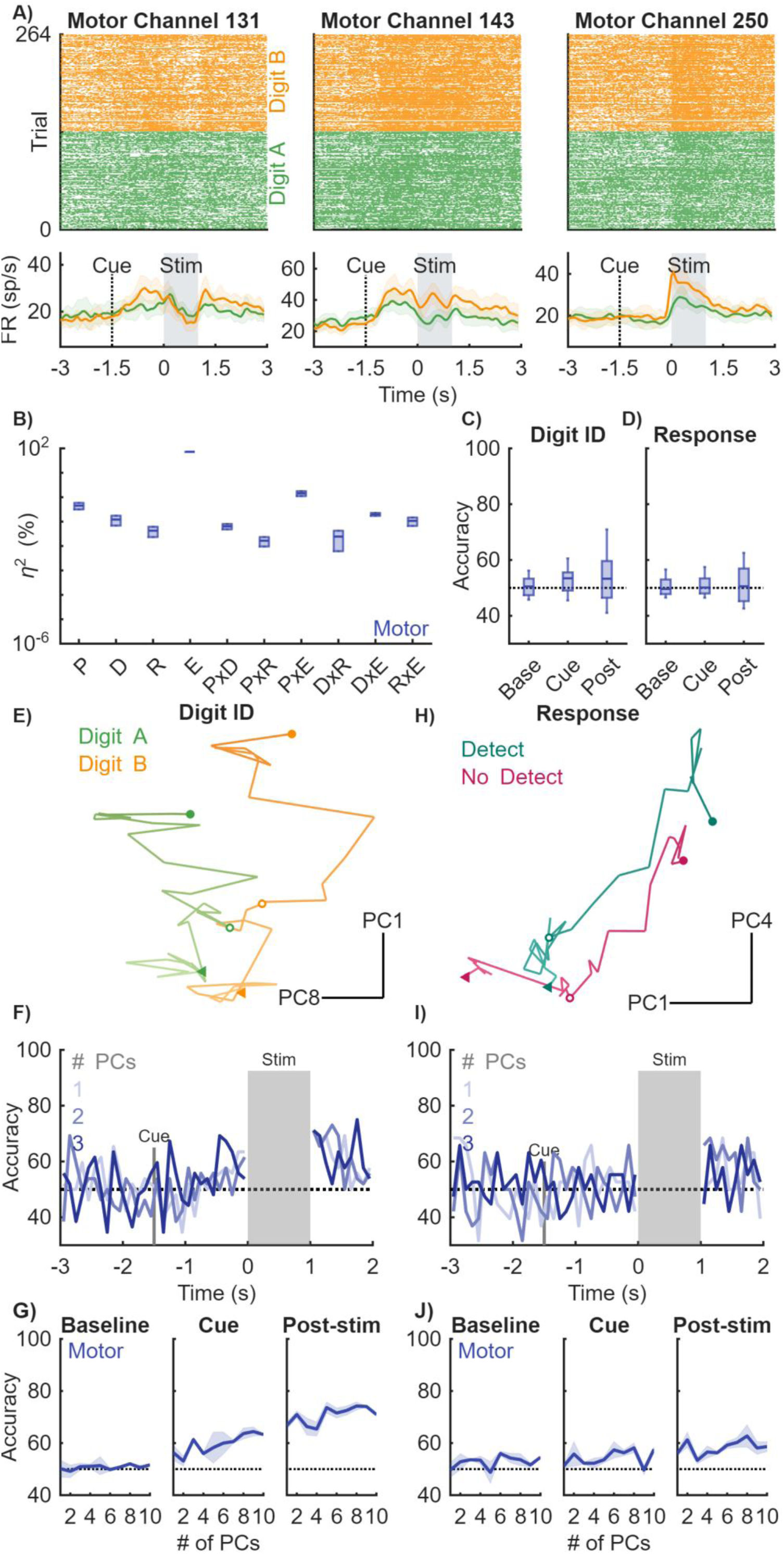
Sensory expectation is consistent across participants. **A)** Rasters (top) and post-stimulus time histograms (bottom) of neural responses throughout the trial. Green and orange indicate cues to different digits. **B)** Variance explained by the period (P) (baseline, cue, post-stim), the digit cued (D), participant’s response (R), and electrode (E). Interaction terms are denoted with a x. **C)** Decoding accuracy for digit ID for the motor array. Dashed line indicated chance classification performance. Boxes indicate 5^th^, 25^th^, 50^th^, 75^th^, and 95^th^ percentiles of variance explained across sessions and channels. **D)** Same as C but for participant response. **E)** Activity from the motor array projected into PC space. Green and orange traces represent the two digits cued. The start of the trial is denoted by a triangle, the cue onset is denoted by an open circle, and the start of stimulation is denoted by a filled in circle. **F)** Decoding of digit identity from the motor array using the first 3 PCs. Dashed line indicates chance performance. **G)** Average decoding performance across sessions as a function of the number of PCs used for the motor array. Performance was grouped into baseline, cue, and post-stimulation epochs by averaging performance during a 1 second interval for baseline and 500ms period for cue and post stimulation. Mean and STD indicated by line and shaded area. Dashed line indicates chance. **H)** Same as E but for participant response. Magenta traces indicate no detect trials while teal traces indicate trials in which the participant reported detecting a stimulus. **I)** Same as F but for participant response. **J)** Same as G but for participant response.

**Supplementary Figure 3.**
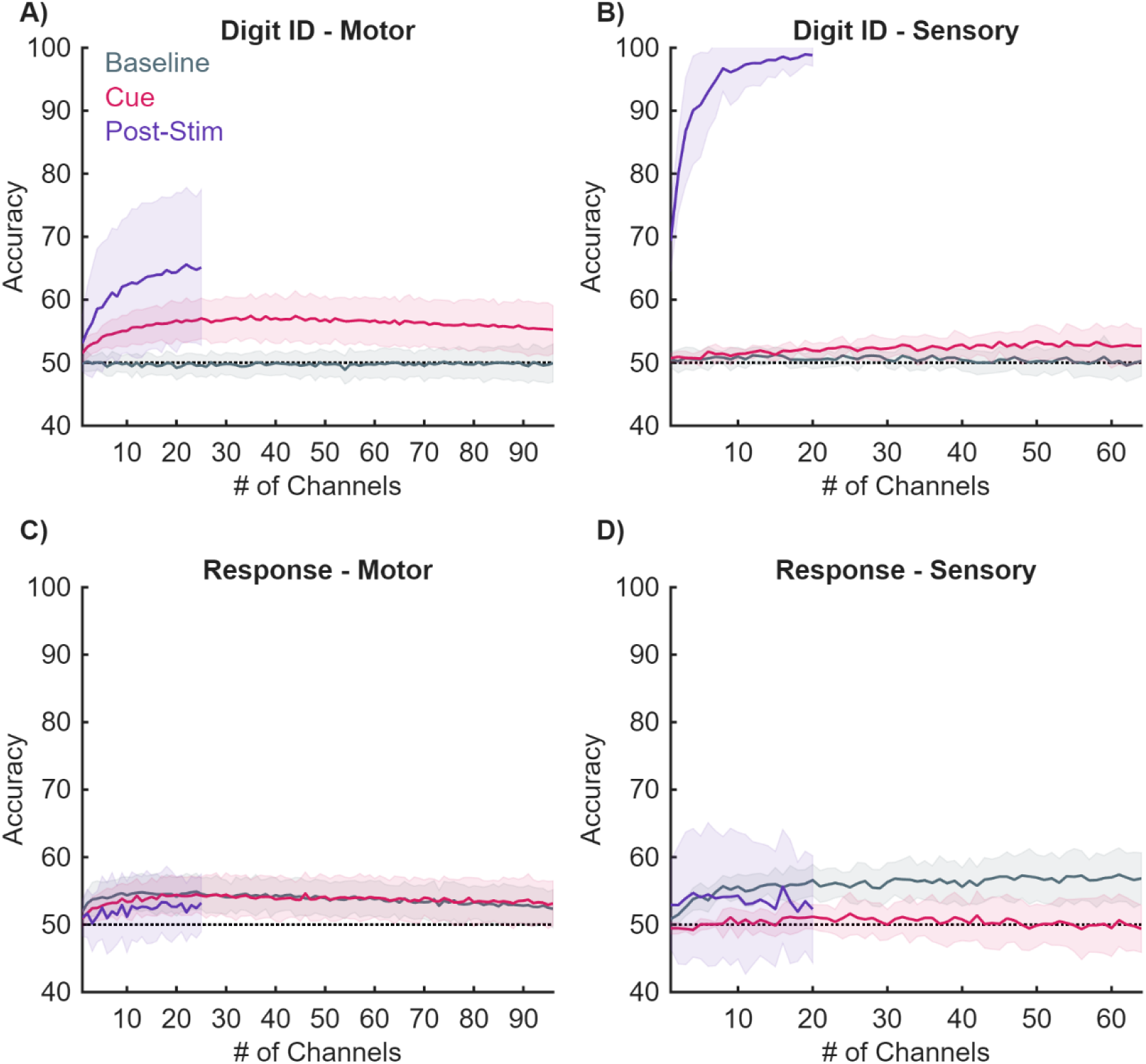
Increasing channel count minimally impacts decoding. **A)** Digit identity was decoded using increasing channel counts (up to 96 channels) from the motor array. Lines indicate mean and STD of bootstrapped classification. Performance plateaus when 10 or more channels are used for decoding. For post-stimulation, channel counts were capped at 25 since there were fewer trials in this condition. **B)** Same as A but for sensory arrays. For post-stimulation, channel counts were capped at 20 since there were fewer trials in this condition. **C)** Same as A but for participant response. **D)** Same as C but for the sensory arrays.

**Supplementary Figure 4.**
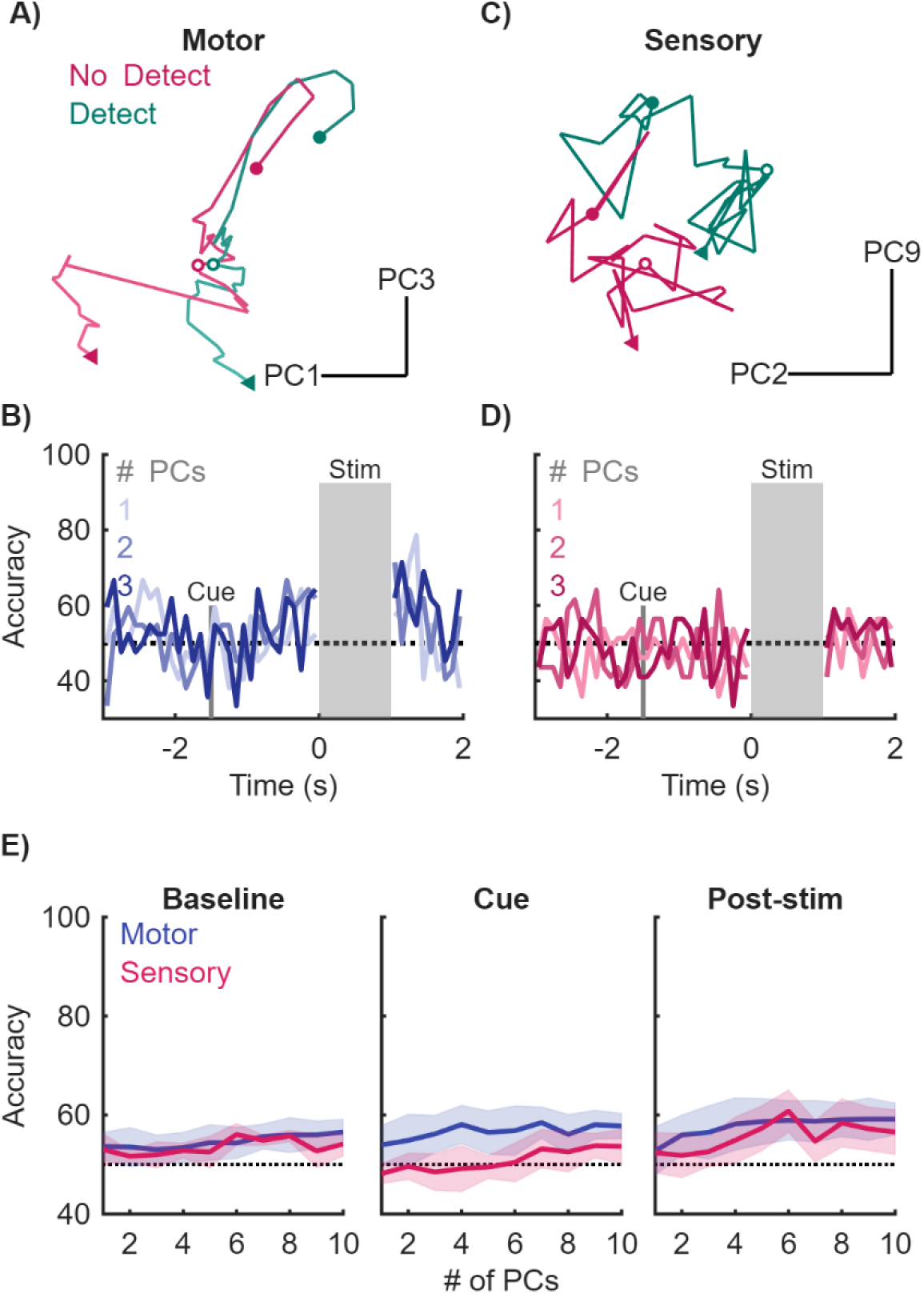
Population responses reflect if the participant detected a stimulus. **A)** Activity from the motor arrays projected into PC space. Magenta traces denote no detect trials while teal traces denote detect trials. The start of the trial is denoted by a triangle, the cue onset is denoted by an open circle, and the start of stimulation is denoted by a filled in circle. **B)** Decoding if a stimulus was detected from the motor array using the first 3 PCs. Performance is at chance level at the start of the trial and increases after the cue is presented and maintained after stimulation. Dashed line indicates chance performance. **C)** Same as A but for the sensory arrays. **D)** The same as B but for the sensory array. **E)** Average decoding performance across sessions as a function of the number of PCs used for the motor (blue) and sensory (pink) arrays. Performance was grouped into baseline, cue, and post-stimulation epochs by averaging performance during a 1 second interval for baseline and 500ms period for cue and post stimulation. Dashed line indicates chance performance and error bars are STD.

**Supplementary Figure 5.**
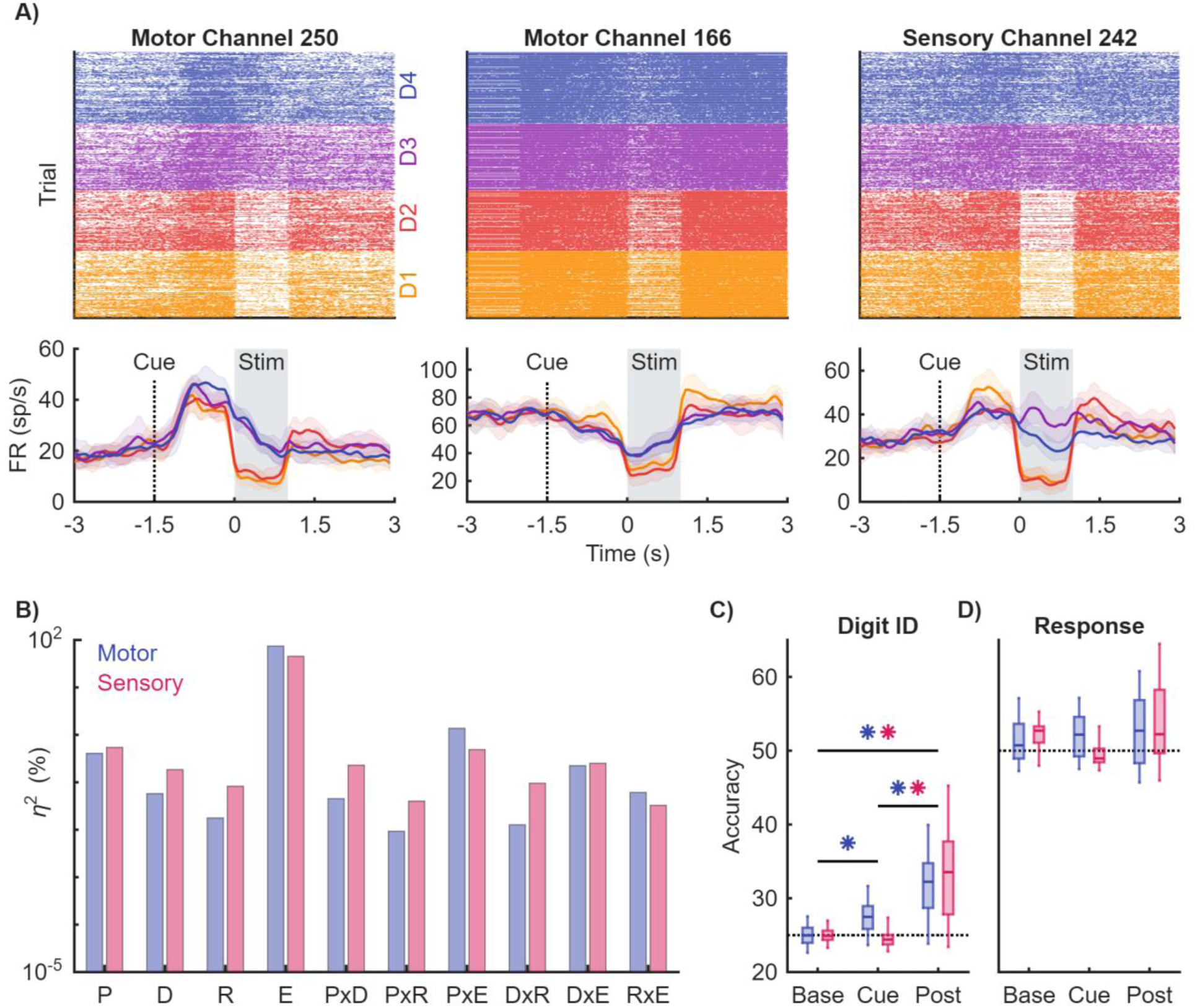
Neural responses to the 4 digit task. **A)** Motor and sensory channels are modulated by the cue and during the post-stimulation period. Rasters (top) and post-stimulus time histograms (bottom) of neural responses throughout the trial. Orange, red, purple and blue colors indicate cues to different digits. **B)** Variance explained by the period (P) (baseline, cue, post-stim), the digit cued (D), participant’s response (R), and electrode (E). Interaction terms are denoted with a x. **C)** Decoding accuracy for digit ID split by motor (blue) and sensory (pink) arrays. Dashed line indicates chance performance. Asterisks denote a significant difference in performance determined by a 1-way ANOVA (p<0.001, Tukey-Kraner post-hoc). **D)** Same as C but for participant response.

**Supplementary Figure 6.**
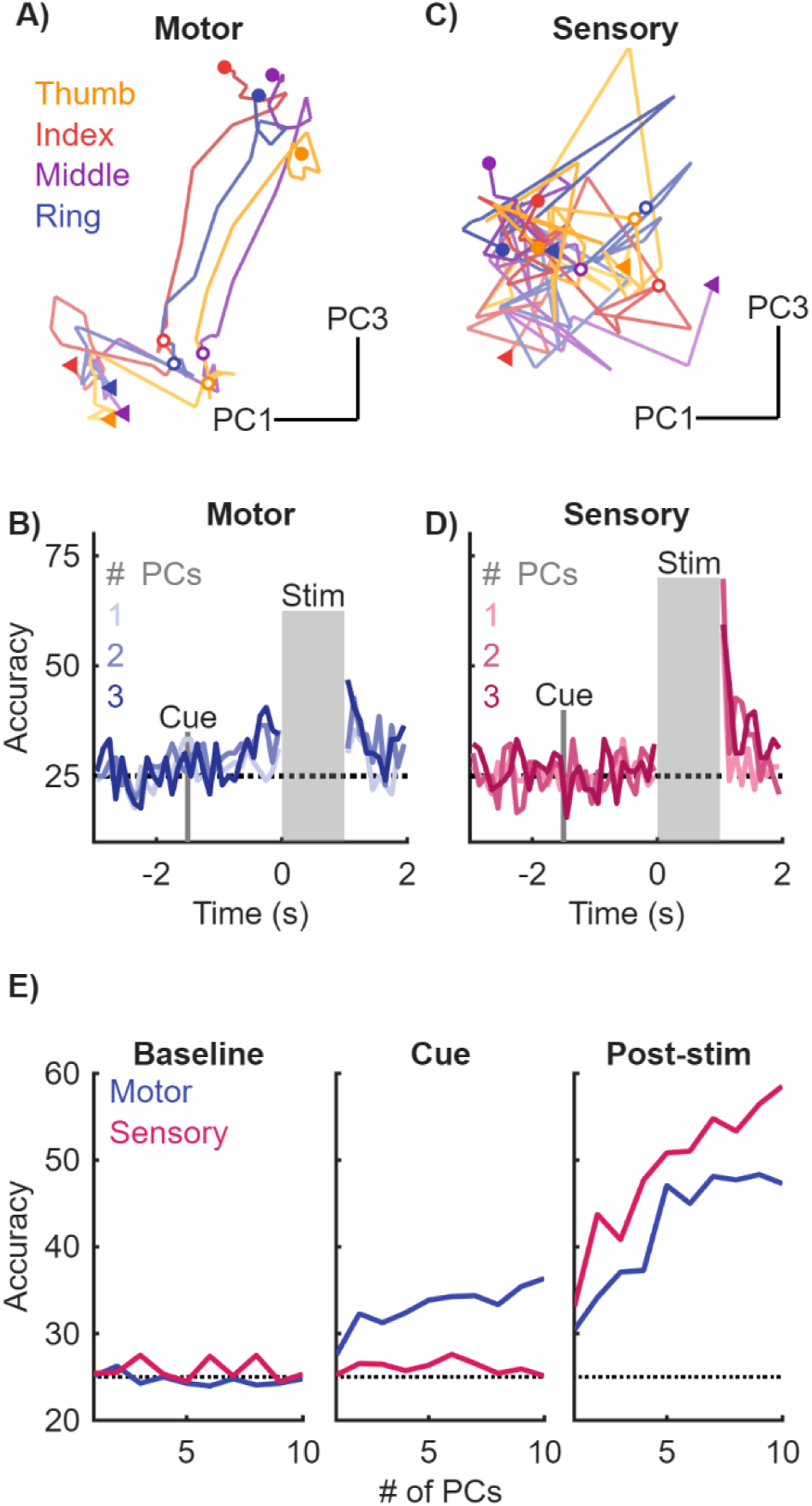
Digit identity decoded from population responses in motor and sensory cortex during the four-digit task. **A)** Activity from the motor array projected into PC space. Different color traces indicate a different digit (thumb – ring). The start of the trial is denoted by a triangle, the cue onset is denoted by an open circle, and the start of stimulation is denoted by a filled in circle. **B)** Decoding of digit identity from the motor array using the first 3 PCs. Performance is at chance level at the start of the trial and increases after the cue is presented and maintained after stimulation. Dashed line indicates chance performance. **D)** The same as A but for the sensory array. **D)** The same as B but for the sensory array. **E)** Average decoding performance as a function of the number of PCs used for the motor (blue) and sensory (pink) arrays. Performance was grouped into baseline, cue, and post-stimulation epochs by averaging performance during a 500ms period at each epoch. Dashed line indicates chance performance (25%).

**Supplementary Figure 7.**
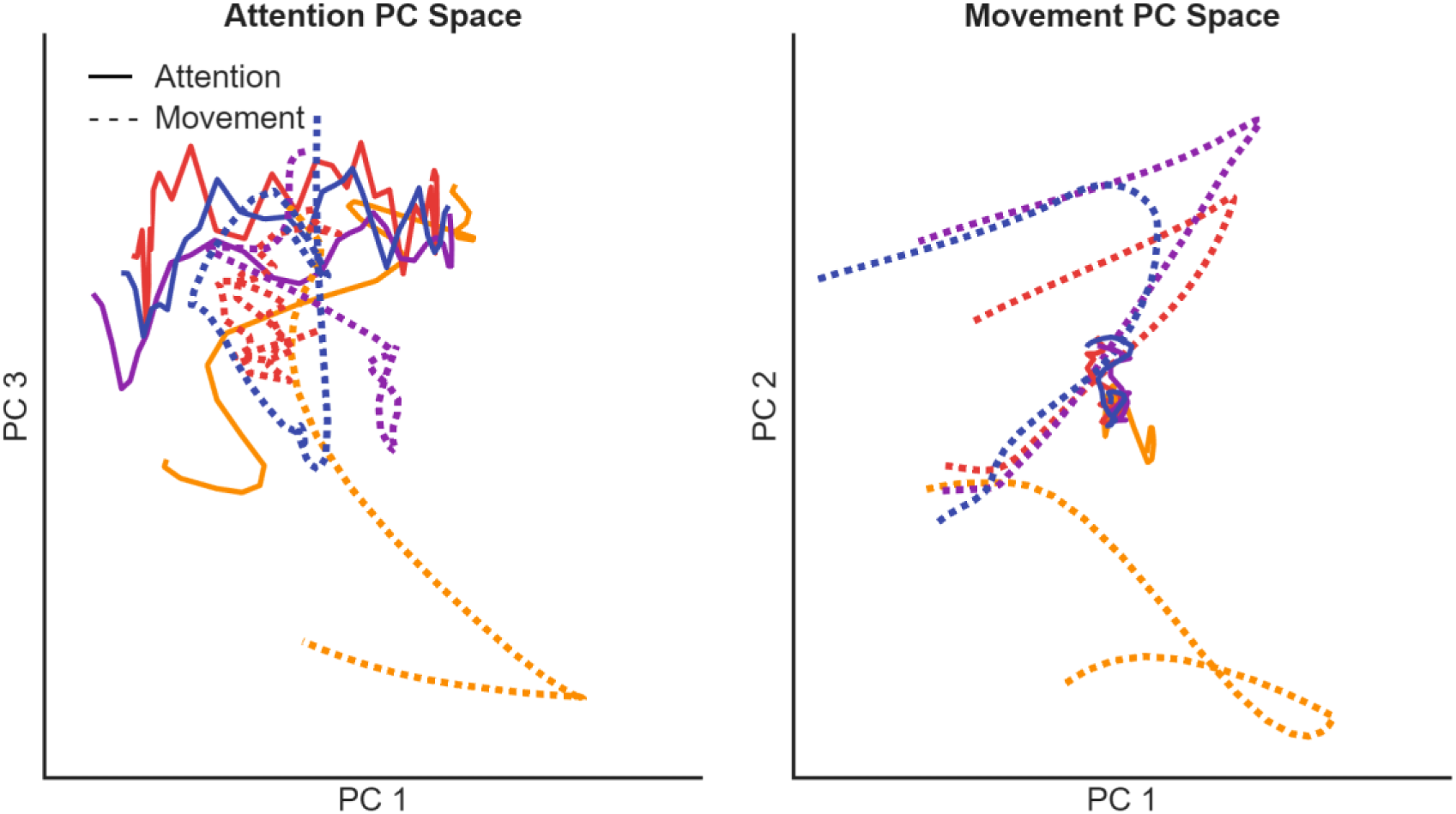
Attention and movement data cross projections. **A)** The attention and movement data projected into attention PC space. Dashed lines indicate movement data and solid lines are attention data. Different colors denote different digits (thumb – ring). **B)** The same as in A but for attention and movement data projected into movement PC space.

**Supplementary Figure 8.**
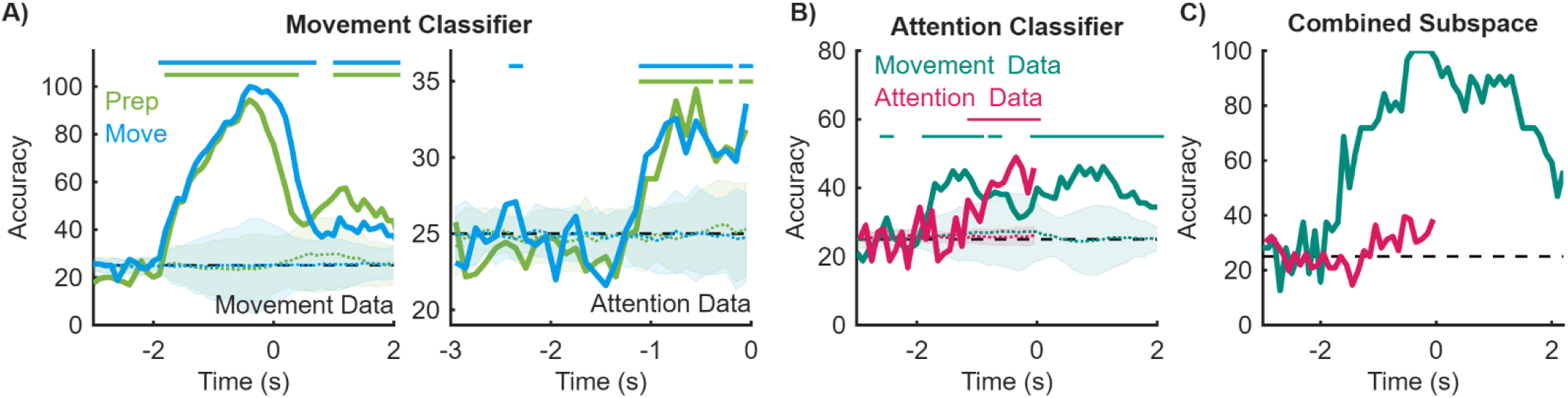
Decoding of digit identity using different models. **A)** Decoding accuracy for digit identity using the movement (left) and attention (right) data as input into a classifier trained on movement data. Green line is the decoding performance when using a classifier trained on the movement preparation period and the blue line indicates performance using a classifier trained on the movement period. Dotted lines represent decoding accuracy on shuffled data, error bars are STD. Horizontal lines indicate significant difference from shuffled data (p ≤ 0.05). Note different y-axes. **B)** Decoding digit identity using a classifier trained on attention data. Teal trace represents decoding accuracy for the movement data and magenta line is the decoding accuracy for the attention data. Dotted lines represent decoding accuracy on shuffled data, error bars are STD. Horizontal lines indicate significant difference from shuffled data (p ≤ 0.05). **C)** Decoding digit identity using data projected into the combined subspace.

